# Long-term effects of caesarean delivery on health and behavioural outcomes of the mother and child in Bangladesh

**DOI:** 10.1101/2020.03.12.20034975

**Authors:** Md. Nuruzzaman Khan, Md. Mostafizur Rahman, Md Aminur Rahman, Mahmudul Alam, Md. Alam Khan

## Abstract

**Background:** Increasing rate of unnecessary caesarean section (CS) is now reported worldwide that intensified the occurrence of adverse health outcomes for mother-child dyads. We investigated the association of CS with some basic health and behaviour outcomes of the mother-child dyads in Bangladesh.

**Methods:** We conducted a community based case-control study from May to August 2019. Total of 600 (300 had CS, and 300 had vaginal delivery (VD)) mother-child dyads were interviewed through a structured questionnaire. Method of delivery was the exposure variable classified as CS and VD. The outcome variables were a group of health and behaviour problems of the mothers and their children. A series of binary logistic regression models were carried out to examine the effects of the exposure variable on outcome variables. Mother’s socio-demographic and reproductive characteristics were adjusted in the models.

**Results:** The mean maternal age (±SD) and weight were 25.1 (±5.2) years and 53.1 (±7.2) kilogram, respectively. Higher likelihood of headache, after delivery hip pain, problem of daily activities, and breastfeeding problem were reported among mother had CS in their last birth than VD. Children born through the CS were reported a higher likelihood of breathing problem and frequent illness and a lower likelihood of food demand and sleeping.

**Conclusion:** The occurrence of CS increases the risk of health and behaviour problems among mothers and their children. This suggests the need for polices to avoid unnecessary CS and to increase awareness of adverse effects of CS. Frequent health checkup following CS is also important.

## Introduction

Caesarean section (CS) is a method of delivery through surgical inclusion of a mother’s abdomen and uterus. Ideally, this is recommended when the life of a mother and/or her child is in risk and would not be saved otherwise. The World Health Organization recommends an upper limit of CS rate at 15% and cautions against rates between 1% and 5% in order to avoid morbidity and mortality [1, 2]. However, there has been an upward trend in global CS rate in the past few decades [3]. More than 18% of the world births are now ended through CS ranging from 6% in the least developed region to 27% in the more developed region [3]. The rates are high as around 35% in Eastern Asia, 38% in Central America, and 32% in North America and Oceania [3, 4]. The risk for this dramatic increase is multifactorial that are varies by countries’ socio-economic condition. However, maternal demographic and reproductive characteristics, including the age of the pregnancy, birth order, and social and cultural factors have all been implicated this trend [5-7]. Professional practice style of doctors or midwives, as well as increasing malpractice (i.e doctors motivate would-be mothers to undergo CS for making money), are recently been addressed as other contributors of rising CS in developing countries [8-10].

There are verities of the advantages and disadvantages of CS use for maternal and child health. The use of CS prevents around 187,000 maternal and 2.9 million neonatal deaths annually worldwide [4, 11]. In the presence of maternal and fetal complications, CS can effectively reduce urinary incontinence and pelvic prolapse [12]. However, unnecessary CS use significantly increases short and long-term health risks for both mothers and children. Infection, haemorrhage, visceral injury, placenta accrete, and placental abruption are some common associated short term risks of CS [13, 14]. The long-term risks of CS are also noticeable worldwide. It increases the risk of asthma up to 12 years and obesity up to 5 years, that were found in a recent systematic review [12]. Moreover, women with CS are reported a higher likelihood of miscarriage, ectopic pregnancy, and stillbirths in a subsequent pregnancy [15-20]. Higher odds of placental accrete, placental abruption, and uterine rupture are also found higher among the mothers with the previous history of CS than VD [18, 21, 22]. However, the effects of CS on maternal and child physical and behaviour health problems following delivery are rarely been investigated. Therefore, in this study an attempt has been made to examine the effects of CS on some basic physical and behaviour health problems of mothers and children through using population level data.

## Methods

### Data

A community-based retrospective case-control study design was employed from May 2019 to August 2019 in the seven communities (village area [16], sub-urban area [3]) of Rajshahi district in Bangladesh. Around 60,000 people live in these areas with a total of 2,560 births in the past three years of the survey date (based on community health workers report), April, 2016-April, 2019). Among of them 600 mothers-child dyads (300 mothers had CS in their last birth and 300 mothers had VD in their last birth) were randomly selected for this study. This sample size was around three times higher than the estimated sample required to make conclusions on the issue we addressed through the standard formula [23]. Face to face interview was conducted through the Bengali version pre-tested questionnaire. Finally, responses were converted to English. Eight Bachelor of Science (B.Sc.) students collected data. The principal investigator (listed as 3^rd^ author in this paper) trained data collectors about the procedure of data collection and checked the completeness of data. He also monitored data collection in the field level.

### Exposure variable

The delivery status of the mothers (CS *vs* VD) for their last birth occurred within three years of survey date was collected and considered as the main exposure variable. Other exposure variables considered were maternal age of birth, type of place of residence, socio-economic status, education (women, husband), children ever born and body mass index. National-level studies in Bangladesh found these variables are important for occurring CS and associated with many other adverse maternal and child health outcomes [7, 24].

### Outcome variables

We considered a range of maternal and child health outcomes. We first asked mothers about their health problems after delivery to survey date. Child data were also collected thereafter by a separate questionnaire. Both questionnaires were open-ended meaning that we did not give any restriction or options on reporting health problems. Mothers’ reported health outcomes were headache (pain in the head or neck), after delivery hip pain (pain in the buttock or lower back), problem of daily activities (problems associated with daily work suck as cooking, walking), suffering physical problem (problems in any organ in the body including eye problem, backbone pain and others), and breastfeeding problem. A variable was also generated by incorporating the types of physical health problems (classified as no problem, eye problem, backbone pain, and others) and included as an outcome variable. Breathing problem (problem during breath), frequent illness (measured by at least once quarterly), behavioural characteristics (classified as obstinate, restless, and quite), food demand (5-6 times in a day for children under 2 years old and 4-5 times a day for the children above 2 years is considered normal), sleeping hours (40% times a day is considered normal) were considered as child health outcomes.

### Statistical Analysis

We used mean and standard deviation to describe participants’ characteristics. The Pearson’s Chi-squared (χ2) test was used to find out the distribution of different maternal and child health outcomes by the method of delivery (CS *vs* VD). Finally, binary logistic regression analysis was used for each of the health outcomes separately adjusted with basic socio-demographic characteristics. All analysis was performed using statistical package for social science (SPSS) version 20.0.

### Ethical consideration

The Institutional Animal, Medical Ethics, Biosafety and Biosecurity Committee (IAMEBBC) of the Institute of Biological Science, University of Rajshahi, Bangladesh has reviewed and approved this study (approval number was 78/318/IAMEBBC/IBSc). Informed verbal consent was obtained from study participants after exploring the objectives of the study. Confidentiality of the participants was ensured as well.

## Results

A total of 600 mother-child dyads were included, of which 300 mothers gave their last birth through CS. The mean age of the respondents was 25.1 years (±5.2) and mean weight was 53.14 kg (±7.2) (Table 1). Most of the respondents were primary educated and reported at least two births. The mean age of marriage was around 16 years and the mean number of antenatal care visit during pregnancy was around 3 times. The self-reported health problems among the mothers and child across delivery status (CS *vs* VD) are presented in Table 2 and 3. We found significant differences in the occurrence of each type of health problem by the delivery status. Adverse mothers and child health outcomes were found much higher among the mothers’ performed CS. We used a series of binary logistic regression model to examine the effects of occurring different maternal and child health outcomes followed by CS (Table 4). We found CS mothers’ reported 3.57 (95% CI, 2.34-4.62) times higher risk of headache and 3.26 times (95% CI, 2.80-4.24) higher risk of hip pain after delivery as compared to the VD mothers. Significant higher risk of daily activities (OR, 2.68, 95% CI, 1.90-3.87) was also found among the CS mothers. When we classified the type of physical problem, we found a higher likelihood of backbone pain (OR, 3.58, 95% CI, 2.12-4.70) following CS than VD. Moreover, CS mothers were reported around 3 times (OR, 3.19, 95% CI, 2.90-4.20) higher likelihood of problems associated with breastfeeding as compared to VD mothers.

**Table 1:** Background characteristics of the respondents.

| Characteristics | Subject (N) | Crude |
| --- | --- | --- |
| Mean (SD) |  |  |
| Maternal age, years | 600 | 25.1 ($\pm 5.2$) |
| Weight (In Kg) | 600 | 53.14 ($\pm 7.2$) |
| Education (years) | 600 | 5.40 ($\pm 2.9$) |
| Children ever born | 600 | 1.9 ($\pm 1.1$) |
| Age at first marriage | 600 | 15.9 ($\pm 3.1$) |
| Number of antenatal visit | 600 | 3.22 ($\pm 1.3$) |

**Table 2:** Physical health problem by delivery status.

| **Physical problem** | **Vaginal delivery**<br>**mothers, %**<br>**(n=300)** | **Caesarean**<br>**delivery**<br>**mothers,%**<br>**(n=300)** | ***p*-value** |
| --- | --- | --- | --- |
| **Headache** |  |  |  |
| Yes | 140 (46.7) | 250 (83.3) | <0.01 |
| No | 160 (53.3) | 50 (16.7) |  |
| **After delivery hip**<br>**pain** |  |  |  |
| Yes | 108 (36.0) | 260 (86.7) | <0.01 |
| No | 192 (64.0) | 40 (13.3) |  |
| **Problem of daily**<br>**activities** |  |  |  |
| Yes | 60 (20.0) | 280 (93.3) | <0.01 |
| No | 240 (80.0) | 20 (6.2) |  |
| **Suffering physical**<br>**problem** |  |  |  |
| Yes | 120 (40.0) | 280 (93.3) | <0.01 |
| No | 180 (60.0) | 20 (6.7) |  |
| **Types of physical**<br>**health problem** |  |  |  |
| **No problem** | 150 (50.0) | 20 (6.7) | <0.01 |
| Eye problem | 50 (16.7) | 70 (23.3) |  |
| Backbone pain | 70 (23.3) | 180 (60.0) |  |
| **Breast feeding**<br>**problem** |  |  |  |
| Yes | 111 (37.0) | 250 (83.0) | <0.01 |
| No | 189 (63.0) | 50 (17.0) |  |

**Table 3:** Child health problem by mother delivery status.

| **Categories** | **Normal delivery**<br>**baby,%**<br>**(N=300)** | **Caesarean delivery**<br>**baby, %**<br>**N=300** | ***p*-value** |
| --- | --- | --- | --- |
| **Breathing problem** |  |  |  |
| Yes | 60 (20.0) | 217 (72.3) | <.01 |
| No | 240 (80.0) | 83 (28.0) |  |
| **Frequent illness** |  |  |  |
| Yes | 52 (17.3) | 230 (76.7) | <.01 |
| No | 248 (82.7) | 70 (23.3) |  |
| **Behavioural**<br>**characteristic** |  |  |  |
| Obstinate | 80 (26.7) | 20 (6.7) | <0.05 |
| Restless | 80 (26.7) | 160 (53.4) |  |
| Quite | 140 (46.7) | 120 (40.0) |  |
| **Food demand** |  |  |  |
| Little | 44 (14.7) | 240 (53.3) | <0.01 |
| Normal | 256 (85.3) | 60 (20.0) |  |
| **Sleeping tendency** |  |  |  |
| Few | 78 (26.0) | 80 (26.7) | <0.01 |
| Normal | 222 (74.0) | 220 (73.3) |  |

**Table 4:** Odds ratio of mothers’ and child health’ problems among mothers who have performed cesarean delivery than vaginal delivery.

| **Women health problem** | **Odds ratio (95% CI)** | **Children health problem** | **Odds ratio (95% CI)** |
| --- | --- | --- | --- |
| **Headache** |  | **Breathing problem** |  |
| Vaginal delivery | 1 | Vaginal delivery | 1 |
| Caesarean delivery | 3.57 (2.34-4.62) | Caesarean delivery | 2.62 (1.90-3.67) |
| **After delivery hip pain** |  | **Frequent illness** |  |
| Vaginal delivery | 1 | Vaginal delivery | 1 |
| Caesarean delivery | 3.26 (2.80-4.24) | Caesarean delivery | 5.10 (3.90-6.20) |
| **Problem of daily activities** |  | **Behavioural characteristics** |  |
| Vaginal delivery | 1 | Obstinate | 1 |
| Caesarean delivery | 2.68 (1.90-3.87) | Restless | 0.45 (0.12-0.90) |
| **Suffering physical problem** |  | Quite | 1.93 (1.01-2.89) |
| Vaginal delivery | 1 | **Food demand** |  |
| Caesarean delivery | 4.15 (2.90-4.12) | Vaginal delivery | 1 |
| **Types of physical health problem** |  | Caesarean delivery | 0.45 (0.12-0.98) |
| No problem |  | **Sleeping tendency** |  |
| Eye Problem | 1.38 (0.95-4.20) | Vaginal delivery | 1 |
| Backbone pain | 3.58 (2.12-4.70) | Caesarean delivery | 0.69 (0.20-1.00) |
| Others | 1.28 (0.90-2.20) |  |  |
| **Breastfeeding problem** |  |  |  |
| Vaginal delivery | 1 |  |  |
| Caesarean delivery | 3.19 (2.90-4.20) |  |  |
| **Constant** | 4.446 | **Constant** | 1.061 |

The children born by CS were reported 2.62 times (95% CI, 1.90-3.67) higher odds of breathing problem after delivery than born by VD. The occurrence of frequent illness was also found around 5 times (OR, 5.10; 95% CI, 3.90-6.20) higher among CS children as compared to the VD children. In addition, children born by CS as compared to VD were reported a higher odds of quite behaviour (OR,1.93, 95% CI, 1.01-2.89). Normal food demand was found 55% (OR, 0.45, 95% CI, 0.12-0.98) lower among the CS children than VD children. The CS children also reported the lower odds of normal sleeping (OR, 0.69, 95% CI, 0.20-1.00) then the children born by VD.

## Discussion

CS is a life-saving procedure which is recommended only when the VD is not possible or would pose the mothers or children in danger. However, a higher number of CS operation is now performed unnecessary which found associated with several adverse mother’ and child’ health outcomes. In this study, we examined the association between CS and some basic health and behaviour problems of the mother and child.

The introduction of CS had been associated with the improvement of maternal and perinatal health outcomes up to 1985 when the rates of CS was around 10-15% [25]. However, when the rates had been crossed 15% in the earlier part of 1990, it becomes concern since increasing rates of CS was showed no evidence of leading to the further improvement of maternal and perinatal health [25, 26]. In addition, CS without any medical indication which is common in the present decades worldwide has been warranted higher maternal and neonatal mortalities, morbidities and complications than does VD [27]. Even at some points, these risk outweighs the potential benefits associated with CS [26]. A 14 years follow up study in Canada of more than 2 million women found 3 times more risk of mortality or complications including blood clots and heart attacks among the mother performed CS unnecessary as compared VD mother [28]. Similarly, higher odds of suffering respiratory distress, and metabolic and immune diseases are reported among children born by CS than VD [28, 29]. The most important concern is that mother and child whose undergo CS need more time to recover and at some points, the problem stays life-long [4, 19, 20, 29].

In this study, we found higher likelihoods of several adverse health and behaviour problems among the mother undergo CS and children who were born by CS. Among CS mother, more than 83% reported headache after delivery and the risk was 3.57 times higher among CS mother than VD mother. This rate was around similar for after delivery hip pain among CS mother. This often a side effect of regional anaesthesia. Before CS doctor inserts a needle in mother spinal area for administrating the pain medication. However, when such insertion punctures cover spinal cord leads to spinal fluids seepage and cause post-surgery headache [30-32]. Most often mothers’ suffering this problem recover within a weeks after delivery [31]. However, in some cases, the problem may stay longer period and could influence women daily activities and other associated physical health problems which were also found higher among CS mother in this study. However, it is unlikely that these problems after CS are common only in developing countries and depend on socio-demographic characteristics. A qualitative study in Australia found almost all CS mothers’ faced problem in daily activities that was hindered by the range of health issues including pain and reduce mobility, abdominal wound problems, infection, infection, vaginal bleeding and urinary incontinence

[33]. We are expecting these problems are more common in Bangladeshi’s CS mother since most of them are not received post-partum care after delivery [34]. We recommended a further broader study to examine sorts of health problems that could contribute negatively to CS mothers’ daily activities. Another problem found in this study was a lower odds of exclusive breastfeeding among CS mothers that are consistent with many other studies in developed and developing countries [35-38]. This might be associated with lower earlier skin contact of mother and child soon after delivery and illness of the mothers and children [39]. Mother earlier return to the formal work should be another predictors since educated and higher socio-economic statuses women usually preferred CS rather than VD [35, 39]. Lower exclusive breastfeeding could also contribute to several adverse health outcomes for children. The major problems associated with breastfeeding duration are frequent illness, child food demand, and nutritional disorder [35]. Researchers of a recent nationally representative study in Bangladesh concluded lowering exclusive breastfeeding associated with increasing infectious diseases of a child including diarrhoea and acute respiratory infection [40]. They also concluded around 28% and 10% of the total occurrence of diarrhoea and acute respiratory infection among children aged 6 months or low in Bangladesh could be possible to reduce by ensuring continues breastfeeding up to six months [40]. However, in this study, we have provided new set evidence regarding the association between use of CS and increasing risk of several health problems including breathing problem, frequent illness, quite behavioural characteristics, lowering food demand and sleeping tendency. Other than lower exclusive breastfeeding poor mother-child bonding among CS, and lower caring of child might associated with these adverse children’ health outcomes though not have been established yet by literature. Further long-term study on targeting of these adverse outcomes among CS mother could be helpful to establish this causal association.

This study has several advantages and some limitations. In our knowledge, this is the first study that considers CS and range of adverse mother and child health and behavioural outcomes. In addition, this study could be helpful for policy-making in Bangladesh as well as other developing countries where CS is now increasing rapidly. We also adjusted our findings with a range of socio-demographic characteristics of the mother and child. However, our relationship is correlational only rather than causal. Moreover, our findings are not nationally representative.

## Conclusion

Use of CS increases the risk of several adverse mother-child health and behavioural outcomes. The risk of headache, after delivery hip pain, the problem of daily activities, suffering physical problem, and breastfeeding problem were found higher among CS mother. In addition, children born by CS reported a higher risk of breathing problem, frequent illness, and quite behavioural characteristics and lower risk of normal food demand and sleeping tendency. Avoiding unnecessary CS and frequent monitoring of the CS’ mothers and children should be considered.

## Data Availability

Data available on request.

## Acknowledgement

We acknowledge the support of the Department of Population Sciences and Human Resource Development, University of Rajshahi, Bangladesh where this study was conducted. We also acknowledge all mothers who were participated in this study.

## Conflict of interest

None

### Abbreviations

CS: Caesarean section
VD: vaginal delivery
OR: odds ratio
95% CI: 95% Confidence Interval

